# Development and Validation of the First FDA Authorized Artificial Intelligence/Machine Learning Diagnostic Tool for the Prediction of Sepsis Risk

**DOI:** 10.1101/2024.05.06.24306954

**Authors:** Akhil Bhargava, Carlos López-Espina, Lee Schmalz, Shah Khan, Gregory L. Watson, Dennys Urdiales, Lincoln Updike, Niko Kurtzman, Alon Dagan, Amanda Doodlesack, Bryan A. Stenson, Deesha Sarma, Eric Reseland, John H. Lee, Max S. Kravitz, Peter S. Antkowiak, Tatyana Shvilkina, Aimee Espinosa, Alexandra Halalau, Carmen Demarco, Francisco Davila, Hugo Davila, Matthew Sims, Nicholas Maddens, Ramona Berghea, Scott Smith, Ashok V. Palagiri, Clinton Ezekiel, Farid Sadaka, Karthik Iyer, Matthew Crisp, Saleem Azad, Vikram Oke, Andrew Friederich, Anwaruddin Syed, Falgun Gosai, Lavneet Chawla, Neil Evans, Kurian Thomas, Roneil Malkani, Roshni Patel, Stockton Mayer, Farhan Ali, Lekshminarayan Raghavakurup, Muleta Tafa, Sahib Singh, Samuel Raouf, Sihai Dave Zhao, Ruoqing Zhu, Rashid Bashir, Bobby Reddy, Nathan I. Shapiro

## Abstract

**Background:** Sepsis is a life-threatening condition that demands prompt treatment for improved patient outcomes. Its heterogenous presentation makes early detection challenging, highlighting the need for effective risk assessment tools. Artificial Intelligence (AI) models have the potential to accurately identify septic patients, but none have previously been FDA-authorized for commercial use. This study outlines the development and validation of the Sepsis ImmunoScore, the first FDA-authorized AI-based software designed to identify patients at risk of sepsis.

**Method:** In this prospective study, adult patients (18+) suspected of infection, as indicated by a blood culture order, were enrolled from five U.S. institutions between April 2017 and July 2022. The participants were divided into an algorithm development cohort (n=2,366), an internal validation cohort (n=393), and an external validation cohort (n=698). The primary endpoint was the presence of sepsis (Sepsis-3) within 24 hours of test initiation. Secondary endpoints included hospital length of stay, ICU admission within 24 hours, mechanical ventilation use within 24 hours, vasopressor use within 24 hours, and in-hospital mortality.

**Results:** The Sepsis ImmunoScore demonstrated high diagnostic accuracy, with an AUC of 0.85 (0.83–0.87) in the derivation cohort, 0.80 (0.74–0.86) in internal validation, and 0.81 (0.77– 0.86) in external validation. The score was categorized into four risk levels for sepsis with corresponding likelihood ratios: low (0.1), medium (0.5), high (2.1), and very high (8.3). These risk categories also predicted in-hospital mortality: low (0.0%), medium (1.9%), high (8.7%), and very high (18.2%) in the external validation cohort. Similar trends were observed for other metrics, such as hospital length of stay, ICU utilization, mechanical ventilation, and vasopressor use.

**Conclusions:** The Sepsis ImmunoScore demonstrated high accuracy for identification and prediction of sepsis and critical illness that could enable prompt identification of patients at high risk of sepsis and adverse outcomes, potentially improving clinical decision-making and patient outcomes.

**Description:** Sepsis is a life-threatening acute condition that requires accurate and rapid identification to guide proper treatment. This study outlines the development and validation of the first FDA-authorized AI-based software to identify patients at risk of having sepsis.

## Introduction

Sepsis is a serious medical condition caused by a dysregulated immune response to infection, which can lead to organ dysfunction and significant morbidity and mortality.^1^ Early treatment, particularly with antibiotics, can improve patient outcomes.^2–7^ However, heterogeneity in the presentation of sepsis makes early recognition difficult, leading to increased mortality.^8^ As a result, there is an opportunity for risk assessment tools to assist clinicians in the quick and accurate identification of patients at high risk of sepsis. Many previously proposed risk assessment tools exist, including clinical approaches, laboratory tests, and sepsis-specific biomarkers; however, none are universally accepted as routine in clinical practice.

To address the need for an informative diagnostic and risk assessment tool in the hospital setting, we developed the Sepsis Immunoscore. The Sepsis ImmunoScore is a tool that uses machine learning to aid in identifying patients likely to have or progress to sepsis within 24 hours of patient assessment. It was granted marketing authorization (De Novo pathway) by the United States Food and Drug Administration (FDA) in April 2024 as the first-ever AI diagnostic authorized for sepsis. The Sepsis ImmunoScore inputs up to 22 parameters derived from patient demographics, vital signs, routinely accepted general clinical laboratory tests, and sepsis specific biomarkers to generate a composite risk score. The risk score categorizes patients into one of four discrete risk groups based on the risk of sepsis within 24-hours. The Sepsis ImmunoScore embeds into a hospital EMR and functions as a diagnostic test, allowing healthcare providers to order and view the test results for a particular patient in the Electronic Health Record (EHR) system, similar to a laboratory test.

In this investigation, we describe the derivation and assess the performance of the Sepsis ImmunoScore functioning as a sepsis risk assessment tool. Accordingly, the objective of this investigation was to evaluate the performance of the Sepsis ImmunoScore and its ability to risk stratify patients for the presence or development of sepsis (defined by Sepsis-3) within 24 hours, and for secondary endpoints of in-hospital mortality, hospital length of stay, ICU admission, mechanical ventilator use, and vasopressor medication use.^9^

## Methods

### Study Design

We conducted a prospective, observational, multi-center study to create a sepsis artificial intelligence/machine learning (AI/ML) algorithm and assess its ability to identify the presence of sepsis within 24 hours, and other secondary outcomes of critical illness morbidity and mortality (**Figure S1**). Participants were enrolled at one of 5 participating hospitals. We obtained study approvals from the ethics boards of participating institutions under a waiver from informed consent, except OSF Saint Francis Medical Center, which required informed consent.

### Study Population

Study inclusion criteria consisted of hospitalized adult patients (aged 18 or older) who had a suspected infection defined by the clinical decision to obtain a blood culture and who had a lithium-heparin (Li-Hep) plasma sample drawn within a 6-hour-window from the first blood culture order that was available for collection. There were no exclusion criteria. Subjects were enrolled between April 2017 and July 2022 from 5 hospital institution sites throughout the United States. The study participants were enrolled in three different cohorts: a *derivation cohort* (n=2,366) where the algorithm was derived, an *internal validation* cohort (n=393) that assessed algorithm performance on a second set of participants from the same hospitals used in the derivation, and a final *external validation* cohort (n=698) that used a new set of participants from hospitals not involved in the algorithm derivation (additional details in supplemental appendix).

### Study Outcomes

#### Endpoints

The primary endpoint was the presence of sepsis at presentation or within 24 hours of study inclusion using the Sepsis-3 criteria: suspected infection and Sequential Organ Failure Assessment (SOFA) score of 2 or greater from baseline.^9^ The derivation cohort used a sepsis-3 outcome derived from the medical record in an automated fashion,^9,10^ while the internal and external validation cohorts used expert clinical adjudication to apply the definitions and determine the sepsis-3 outcome. The clinical adjudication occurred in a retrospective fashion by a clinician who likely did not treat the patient, but had access to the entire hospital chart and utilized information including laboratory testing, radiology testing, clinical assessment and decision-making documentation. The secondary endpoints consisted of sepsis-related metrics of critical illness including: in-hospital mortality, hospital length of stay, ICU admission, use of mechanical ventilator, and use of vasopressors.

### Data Collection

Data were gathered directly through an offline EMR extraction and a transfer of de-identified data that were linked to corresponding patient blood specimens. Data elements were abstracted from the EMR and included demographic information, coded ICD-10 diagnoses, medications, vital sign measurements, clinical laboratory test results (e.g., chemistry laboratory testing results, lactic acid), and sepsis-related laboratory measurements (C-reactive protein and procalcitonin – tested at external lab – see supplemental appendix for details), secondary outcomes metrics, and relevant data to conduct adjudication (e.g., microbiology results), and relevant orders (e.g., antibiotic administration). Comorbidities were based on the components of the Charlson Comorbidity Index (CCI) and were encoded based on ICD-10-CM encodings defined by the National Cancer Institute (NCI) Comorbidity Index/SEER^11^. Immunocompromised patients were identified based on ICD-10-CM encodings defined by Agency for Healthcare Research and Quality (AHRQ).^12^

### Sepsis ImmunoScore

#### Algorithm Development

The Sepsis ImmunoScore machine learning algorithm was created using a supervised, calibrated random forest that predicts the probability of a patient meeting Sepsis-3 criteria within 24 hours of study entry. A random forest was trained on the 2,366 patient encounters in the derivation cohort using 22 patient-specific features comprising demographics, vital signs, and laboratory tests measured close to study entry. Model parameters were optimized using 3 repeats of 5-fold-cross-validation, and missing data were imputed using bagged trees. Predictions were calibrated to the probability of sepsis-3 to compute a sepsis risk score by regressing the outcome on the out-of-bag predictions of the random forest in the derivation cohort.^13^ Sepsis risk scores were divided into four risk stratification categories by thresholds identified during the development process using out-of-bag predictions in the derivation cohort. (See supplemental appendix) Interventional Shapley Additive exPlanation (SHAP) values were calculated for each patient using the out-of-bag measurements to assess for the feature importance of the random-forest model.^13–16^

### Risk Score and Risk Category Generation

To assess performance, the Sepsis ImmunoScore was calculated for patients in the internal and external validation cohorts. Calibrated out-of-bag scores were used for the derivation cohort to reduce bias from overfitting in performance estimation. No result was generated for patients lacking a measurement for procalcitonin (PCT), C-reactive protein (CRP), white blood cell count, platelet count, creatinine, or blood urea nitrogen between 24 hours prior to study entry (blood culture order) and 3.5 hours after. Similarly, no result was generated for patients without a measurement for systolic blood pressure, diastolic blood pressure, inspired oxygen percentage, heart rate, or respiratory rate between six hours prior to study entry and 3.5 hours after. Missing values for the remaining 10 input parameters were imputed by the Sepsis ImmunoScore to produce a sepsis risk score.

### Statistical Analysis

Diagnostic accuracy was assessed by determining the ability of the sepsis ImmunoScore and its corresponding risk stratification category (low, medium, high, or very high), to identify patients with the primary outcome of sepsis (sepsis-3 within 24 hours of study entry) and secondary outcomes. We estimated stratum specific likelihood ratios (SSLR) and predictive values (PV) each of the risk categories and assessed for a monotonic increasing relationship between risk category severity and outcomes using a one-sided Cochran-Armitage hypothesis test.^17–19^ We also estimated the area under the receiver operating characteristic curve (AUROC) of the sepsis risk score. We report all uncertainty intervals using 95% confidence intervals. Confidence intervals for the AUROC were estimated using a binormal approximation estimator for the standard error.^20^ Analyses were conducted using R statistical software version 4.2.1.

#### Sepsis-3 Diagnostic and Prognostic Sensitivity Analysis

Our a priori primary outcome was the presence of sepsis within 24 hours of study inclusion using the Sepsis-3 criteria as described above. This included both patients who met sepsis criteria at initial evaluation as well as those who developed sepsis over the subsequent 24 hours. This decision (made in consultation with the FDA) was driven by the rationale that the treatment is similar for both of these clinically relevant groups (e.g., antibiotics and supportive care). We also performed a sensitivity analysis to assess the ability of the Sepsis Immunoscore to discriminate each of these groups from those patients who never had nor developed sepsis within 24-hours. Specifically, we assessed Sepsis Immunoscore performance separately for: a) ***diagnosis*** of sepsis-3 - defined as the presence of sepsis-3 present at initial evaluation, and the b) **prognosis** of sepsis - defined as the development of sepsis-3 criteria within 24 hours in those who did not have sepsis at presentation. The AUC and stratum specific likelihood ratios were reported for these analyses.

### Sample Size Calculation

This study was powered based on the confidence interval of the AUROC for the sepsis endpoint.^20^ The calculation assumed a sepsis prevalence of 32%, an estimated AUROC of 0.75, a maximum allowable difference between the true AUC and its estimate of 0.023, and a significance level of 0.05 resulting in an estimated sample size of 735 subjects. Additional participants were enrolled beyond these calculations to include participants of varying age, racial backgrounds, ethnicities, and geographic location. The initial study design used a single validation cohort partially enrolled from hospitals included in the derivation set; however, based on direction from the FDA, we split the cohort into the current internal and external validation format.

## Results

There were a total of 3,457 patient encounters included with valid Sepsis ImmunoScore results, with 2,366 encounters in the derivation set, 393 in the internal validation set, and 698 in the external validation set **(Figure S1)**. The study enrolled participants with age, sex, race, and ethnicity, and comorbidities typical of sepsis patients in the U.S. (**Table 1**). The rate of sepsis was 32% in the derivation, 28% in internal validation, and 22% in the external validation cohorts (**Table 1**). Patients with sepsis had higher rates of severe illness and mortality compared to those without sepsis (**Table 1**).

**Table 1.** Baseline Data and Adverse Outcomes for Derivation, Internal Validation and External Validation.

| Characteristic | Patients Encounters, No. (%) |  |  |
| --- | --- | --- | --- |
|  | Derivation | Internal Validation | External Validation |
|  | (N = 2366) | (N = 393) | (N = 698) |
| Clinical Site |  |  |  |
| Beth Israel Deaconess Medical Center – Boston, MA | 0 (0.0) | 0 (0.0) | 356 (51.0) |
| OSF – Peoria, IL | 712 (30.1) | 87 (22.1) | 0 (0.0) |
| Jesse Brown VA - Chicago, IL | 0 (0.0) | 0 (0.0) | 65 (9.3) |
| Mercy Health - St. Louis, MO | 1061 (44.8) | 306 (77.9) | 0 (0.0) |
| Beaumont - Royal Oak, MI | 0 (0.0) | 0 (0.0) | 277 (39.7) |
| Carle Foundation Hospital – Urbana, IL | 593 (25.1) | 0 (0.0) | 0 (0.0) |
| Age (mean (S.D.)) | 64.20 (16.59) | 64.06 (17.66) | 62.80 (17.01) |
| Male (%) | 1195 (50.5) | 210 (53.4) | 391 (56.0) |
| Race |  |  |  |
| American Indian or Alaska Native | 1 (0.0) | 0 (0.0) | 2 (0.3) |
| Asian | 12 (0.5) | 2 (0.5) | 14 (2.0) |
| Black or African American | 315 (13.3) | 57 (14.5) | 154 (22.1) |
| Native Hawaiian or Other Pacific Islander | 0 (0.0) | 0 (0.0) | 1 (0.1) |
| Unknown | 85 (3.6) | 12 (3.1) | 119 (17.0) |
| White | 1953 (82.5) | 322 (81.9) | 408 (58.5) |
| Ethnicity |  |  |  |
| Hispanic or Latino | 26 (1.1) | 2 (0.5) | 96 (13.8) |
| Not Hispanic or Latino | 1725 (72.9) | 385 (98.0) | 564 (80.8) |
| Unknown | 615 (26.0) | 6 (1.5) | 38 (5.4) |
| High-Risk Comorbidities |  |  |  |
| Acute Myocardial Infarction (%) | 97 (4.1) | 11 (2.8) | 43 (6.2) |
| History of Myocardial Infarction (%) | 101 (4.3) | 21 (5.3) | 54 (7.7) |
| Congestive Heart Failure (%) | 583 (24.6) | 103 (26.2) | 170 (24.4) |
| Peripheral Vascular Disease (%) | 225 (9.5) | 49 (12.5) | 72 (10.3) |
| Cerebrovascular Disease (%) | 130 (5.5) | 38 (9.7) | 65 (9.3) |
| Chronic Obstructive Pulmonary Disease (%) | 606 (25.6) | 107 (27.2) | 171 (24.5) |
| Dementia (%) | 167 (7.1) | 45 (11.5) | 72 (10.3) |
| Paralysis (%) | 68 (2.9) | 10 (2.5) | 20 (2.9) |
| Diabetes (%) | 630 (26.6) | 101 (25.7) | 156 (22.3) |
| Diabetes with Complications (%) | 423 (17.9) | 93 (23.7) | 155 (22.2) |
| Renal Disease (%) | 659 (27.9) | 123 (31.3) | 216 (30.9) |
| Mild Liver Disease (%) | 118 (5.0) | 19 (4.8) | 94 (13.5) |
| Moderate and Severe Liver Disease (%) | 45 (1.9) | 6 (1.5) | 55 (7.9) |
| Peptic Ulcer Disease (%) | 45 (1.9) | 8 (2.0) | 13 (1.9) |
| Rheumatologic Disease (%) | 105 (4.4) | 17 (4.3) | 33 (4.7) |
| AIDS (%) | 17 (0.7) | 2 (0.5) | 6 (0.9) |
| Immunocompromised (%) | 470 (19.9) | 117 (29.8) | 190 (27.2) |
| COVID-19 (%) | 189 (8.0) | 28 (7.1) | 73 (10.5) |
| Adverse Outcomes |  |  |  |
| Sepsis-3 within 24 hours (%) | 763 (32.2) | 108 (27.5) | 151 (21.6) |
| In-hospital Mortality (%) | 147 (6.2) | 33 (8.4) | 33 (4.7) |
| ICU Transfer (%) | 491 (27.7) | 144 (36.6) | 151 (21.6) |
| Placement of Mechanical Ventilation (%) | 191 (8.1) | 44 (11.2) | 51 (7.3) |
| Administration of Vasopressors (%) | 223 (9.4) | 53 (13.5) | 77 (11.0) |
| Length of Stay (median [IQR]) | 4.7 [2.6, 8.5] | 4.98 [2.8, 10.8] | 5.94 [3.3, 10.4] |

### Sepsis ImmunoScore Algorithm Development

The Sepsis ImmunoScore algorithm uses up to 22 input parameters to generate the risk score and place patients in one of four discrete risk stratification categories (**Table 2**). The 22 input parameters consist of demographic data (age), vital sign measurements, complete-metabolic-panel measurements, complete blood count panel measurements, lactate, and sepsis biomarkers PCT and CRP. The Interventional SHAP values indicated that the three most influential parameters to the model were PCT, respiratory rate, and systolic blood-pressure (**Figure 1**). The AUC in the derivation set was 0.85 (95% confidence interval: 0.83–0.87) for the medical record derived sepsis outcome (**Table S1**). Additionally, the Sepsis ImmunoScore risk categories were associated with increasing risk of sepsis in the derivation set (**Figure 1, Table S3**).

**Figure 1.**
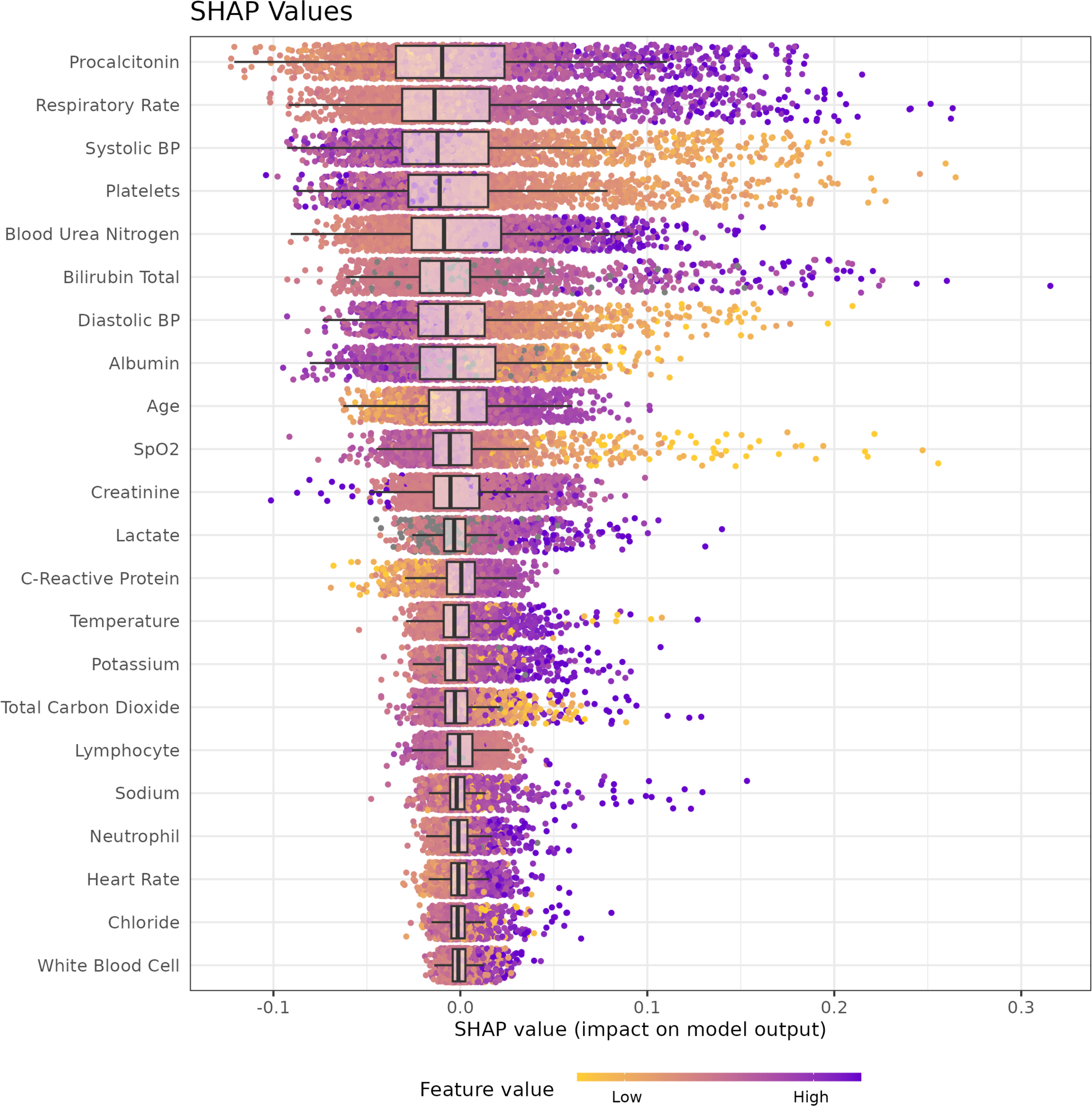
Sepsis ImmunoScore Feature Importance. Scatter plots of the Interventional SHAP values (a measure of feature contribution to the algorithm’s output) for each input feature across all patients in the derivation cohort. The features are listed on y-axis in descending order of importance with the features contributing the most at the top of the list, determined by the mean absolute Interventional SHAP value across patients. Each data point is colored according to its observed standardized measurement value: purple indicates elevated measurements, while yellow indicates lower measurement. The individual values colored in purple offer the most impactful contributions (e.g. high procalcitonin and respirate rate values, and low systolic blood pressure and platelet values all offered substantial contributions) Grey data points represent parameters that were imputed during the generation of the Sepsis ImmunoScore result.

**Table 2.**
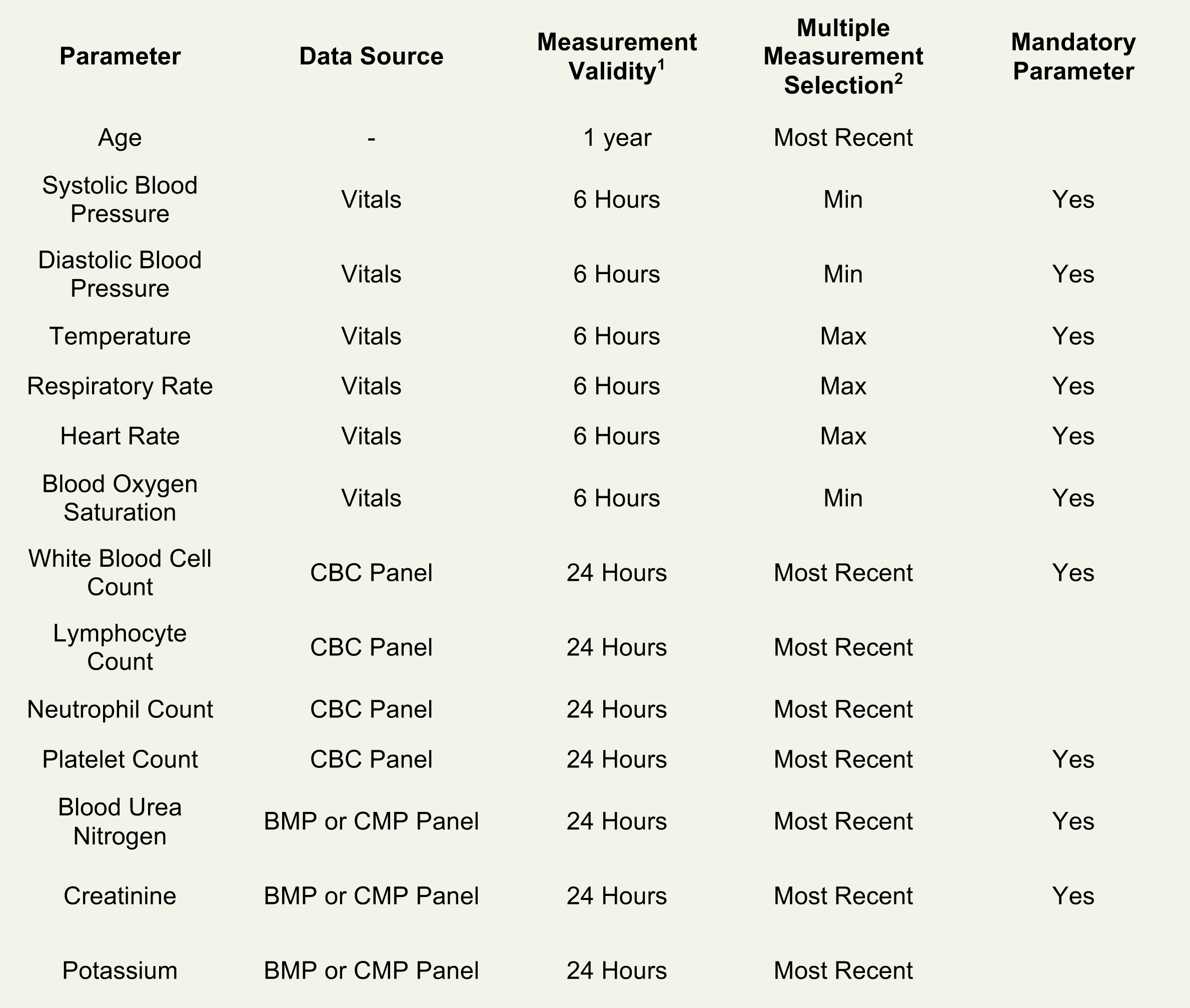
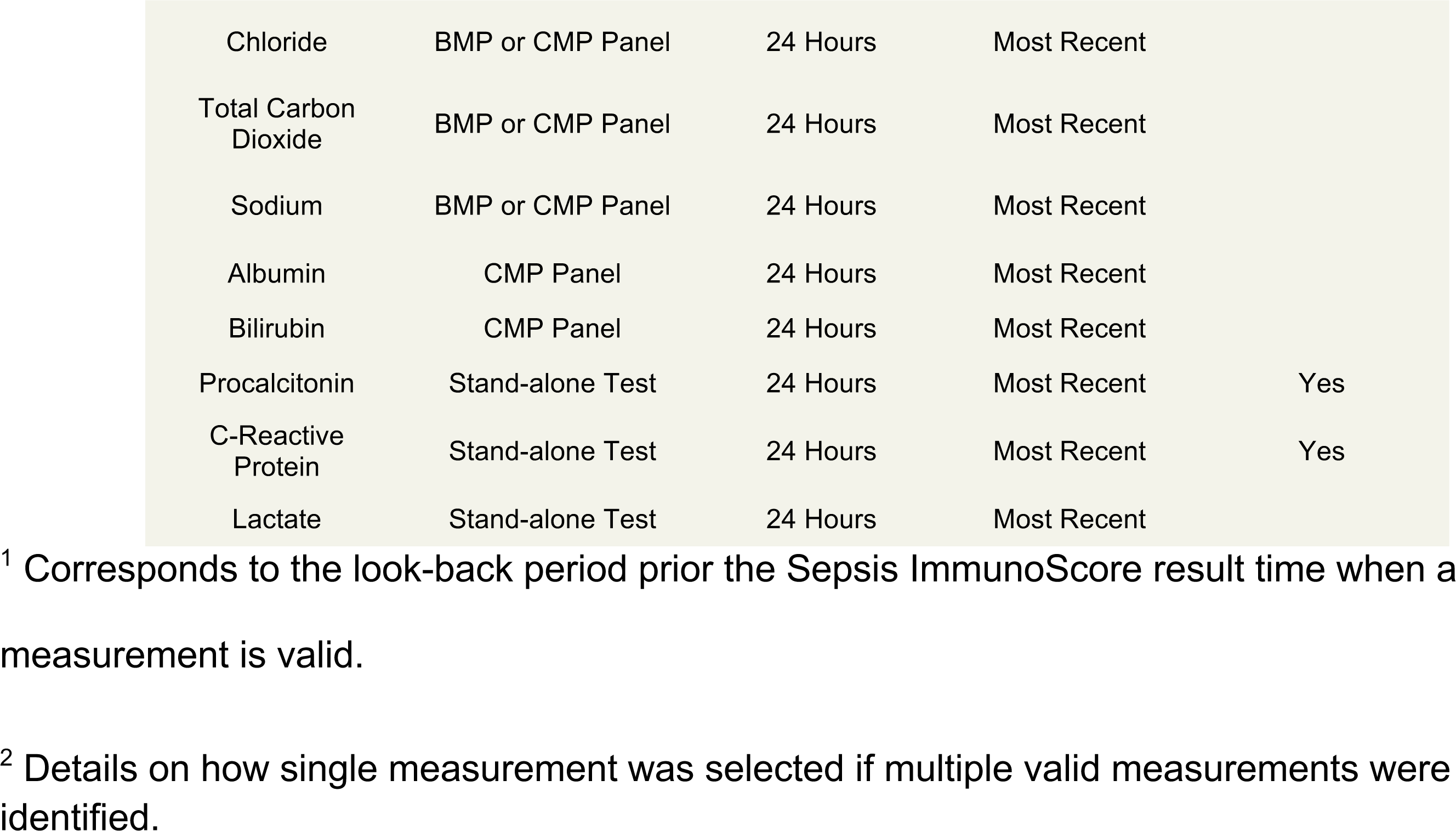
Sepsis ImmunoScore Input Parameters.

| Parameter | Data Source | Measurement Validity<sup>1</sup> | Multiple Measurement Selection<sup>2</sup> | Mandatory Parameter |
| --- | --- | --- | --- | --- |
| Age | - | 1 year | Most Recent |  |
| Systolic Blood Pressure | Vitals | 6 Hours | Min | Yes |
| Diastolic Blood Pressure | Vitals | 6 Hours | Min | Yes |
| Temperature | Vitals | 6 Hours | Max | Yes |
| Respiratory Rate | Vitals | 6 Hours | Max | Yes |
| Heart Rate | Vitals | 6 Hours | Max | Yes |
| Blood Oxygen Saturation | Vitals | 6 Hours | Min | Yes |
| White Blood Cell Count | CBC Panel | 24 Hours | Most Recent | Yes |
| Lymphocyte Count | CBC Panel | 24 Hours | Most Recent |  |
| Neutrophil Count | CBC Panel | 24 Hours | Most Recent |  |
| Platelet Count | CBC Panel | 24 Hours | Most Recent | Yes |
| Blood Urea Nitrogen | BMP or CMP Panel | 24 Hours | Most Recent | Yes |
| Creatinine | BMP or CMP Panel | 24 Hours | Most Recent | Yes |
| Potassium | BMP or CMP Panel | 24 Hours | Most Recent |  |
| Chloride | BMP or CMP Panel | 24 Hours | Most Recent |  |
| Total Carbon Dioxide | BMP or CMP Panel | 24 Hours | Most Recent |  |
| Sodium | BMP or CMP Panel | 24 Hours | Most Recent |  |
| Albumin | CMP Panel | 24 Hours | Most Recent |  |
| Bilirubin | CMP Panel | 24 Hours | Most Recent |  |
| Procalcitonin | Stand-alone Test | 24 Hours | Most Recent | Yes |
| C-Reactive Protein | Stand-alone Test | 24 Hours | Most Recent | Yes |
| Lactate | Stand-alone Test | 24 Hours | Most Recent |  |

<sup>1</sup> Corresponds to the look-back period prior the Sepsis ImmunoScore result time when a measurement is valid.
<sup>2</sup> Details on how single measurement was selected if multiple valid measurements were identified.

### Primary Endpoint

The Sepsis ImmunoScore demonstrated high overall diagnostic accuracy for predicting sepsis, with an AUC in the derivation set of 0.85 (95% confidence interval: 0.83–0.87) for the medical record derived sepsis outcome, and] 0.80 (0.74–0.86) in the internal validation and 0.81 (0.77– 0.86) in the external validation for the adjudicated sepsis outcome (**Table S1**). The Sepsis ImmunoScore risk categories were associated with increasing risk of sepsis in both validation sets (**Figure 2**, **Table 3**, **Table S2**). Of note, in the external validation set, the likelihood ratios were: low 0.1 (0.1–0.2), medium 0.5 (0.3–0.8), high 2.1 (1.8–2.5), very high 8.3 (4.1–17.1) for the primary Sepsis-3. These ratios are monotonically increasing with no overlapping confidence intervals, suggesting stepwise risk discrimination for sepsis.

**Figure 2.**
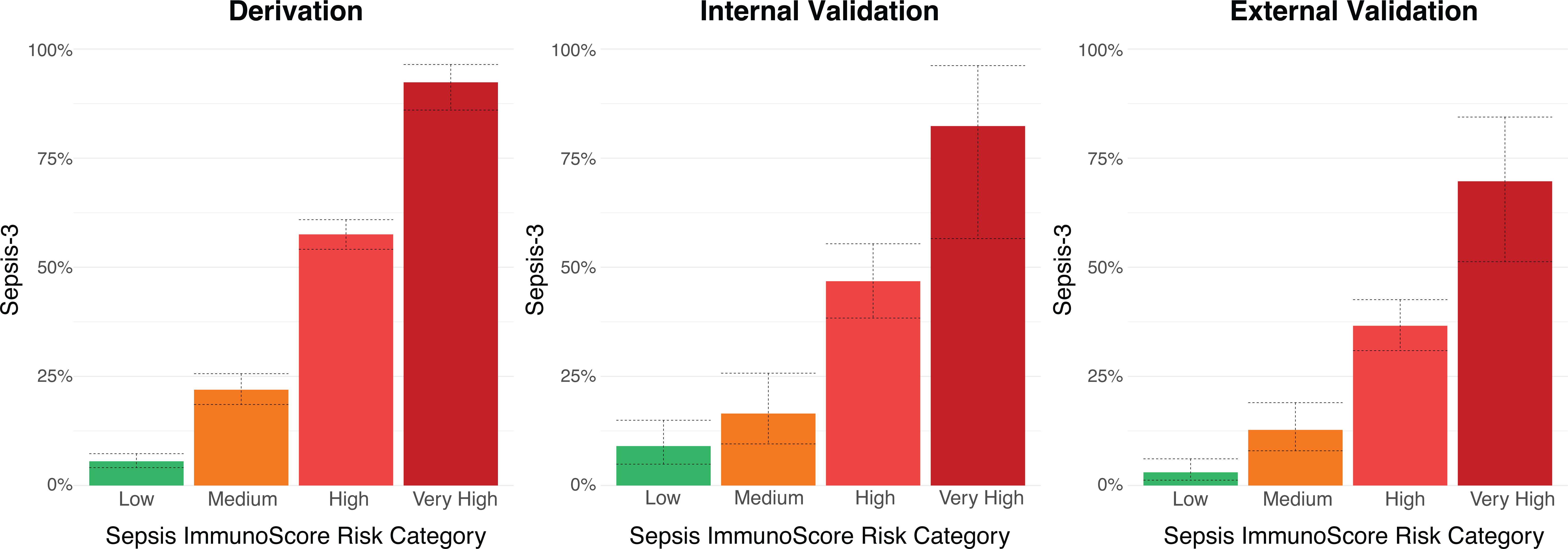
Sepsis ImmunoScore Stratification for Sepsis-3 in all Cohorts. Barplots are shown for the derivation, internal validation, and external validation datasets for the Sepsis-3 within 24 hours P.V.s for each Sepsis ImmunoScore risk stratification category. Dashed lines indicate the 95% C.I.s.

**Table 3.** Sepsis ImmunoScore Risk Stratification for Sepsis-3 Within 24 Hours.

| Cohort | ImmunoScore Risk Category | Patients | Septic Patients | Sepsis P.V. [95% CI] | Sepsis Likelihood Ratio [95% CI] | Cochran-Armitage Test (p-value) |
| --- | --- | --- | --- | --- | --- | --- |
| External | Low | 232 | 7 | 3.0% [1.2%, 6.1%] | 0.1 [0.1, 0.2] | < 0.001 |
| Validation | Medium | 157 | 20 | 12.7% [7.96%, 19.0%] | 0.5 [0.3, 0.8] |  |
| (N = 698) | High | 276 | 101 | 36.6% [30.1%, 42.6%] | 2.1 [1.8, 2.5] |  |
|  | Very High | 33 | 23 | 69.7% [51.3%, 84.4%] | 8.3 [4.1, 17.1] |  |

### Secondary Endpoints

We assessed the ability for the Sepsis ImmunoScore to predict the secondary outcomes of ICU admission within 24 hours, in-hospital mortality, use of mechanical ventilation within 24 hours, and use of vasopressors within 24 hours. The Sepsis ImmunoScore was highly predictive of these outcomes. The Sepsis ImmunoScore categories ranging from low, medium, high, and very high demonstrated good predictive ability based on both rate of outcome as well as the corresponding stratum specific likelihood ratios (**Figure 3**, **Table 4**, **Table S3**). In the external validation cohort, the observed in-hospital mortality rates in the low, medium, high, and very high-risk groups were 0.0% (0.0%, 1.6%), 1.9% (0.40%–5.5%), 8.7% (5.7%–12.7%), and 18.2% (7.0%–35.5%) respectively. Additionally, the observed median number of days for the composite length of stay endpoint in the low, medium, high, and very high-risk groups were: 4.0 (3.5–4.9), 5.7 (4.9–7.0), 7.7 (6.5–8.5), and 13.5 (7.1–19.1) respectively. The proportion of patients transferred to the ICU within 24 hours was 4.7% (2.4%–8.3%), 12.7% (8.0%–19.0%), 25.7% (20.7%–31.3%), and 54.6% (36.4%–71.9%) respectively. Similar trends were observed for mechanical ventilation and vasopressor usage. Cochran-Armitage hypothesis tests indicated statistically significant monotonic increasing relationships between outcome predictive value and risk stratification category severity for each secondary endpoint (*p*-value < 0.01, **Table 4, Table S3**). Risk stratification category severity was also associated with time to event for each secondary endpoint (**Figure S2**).

**Figure 3.**
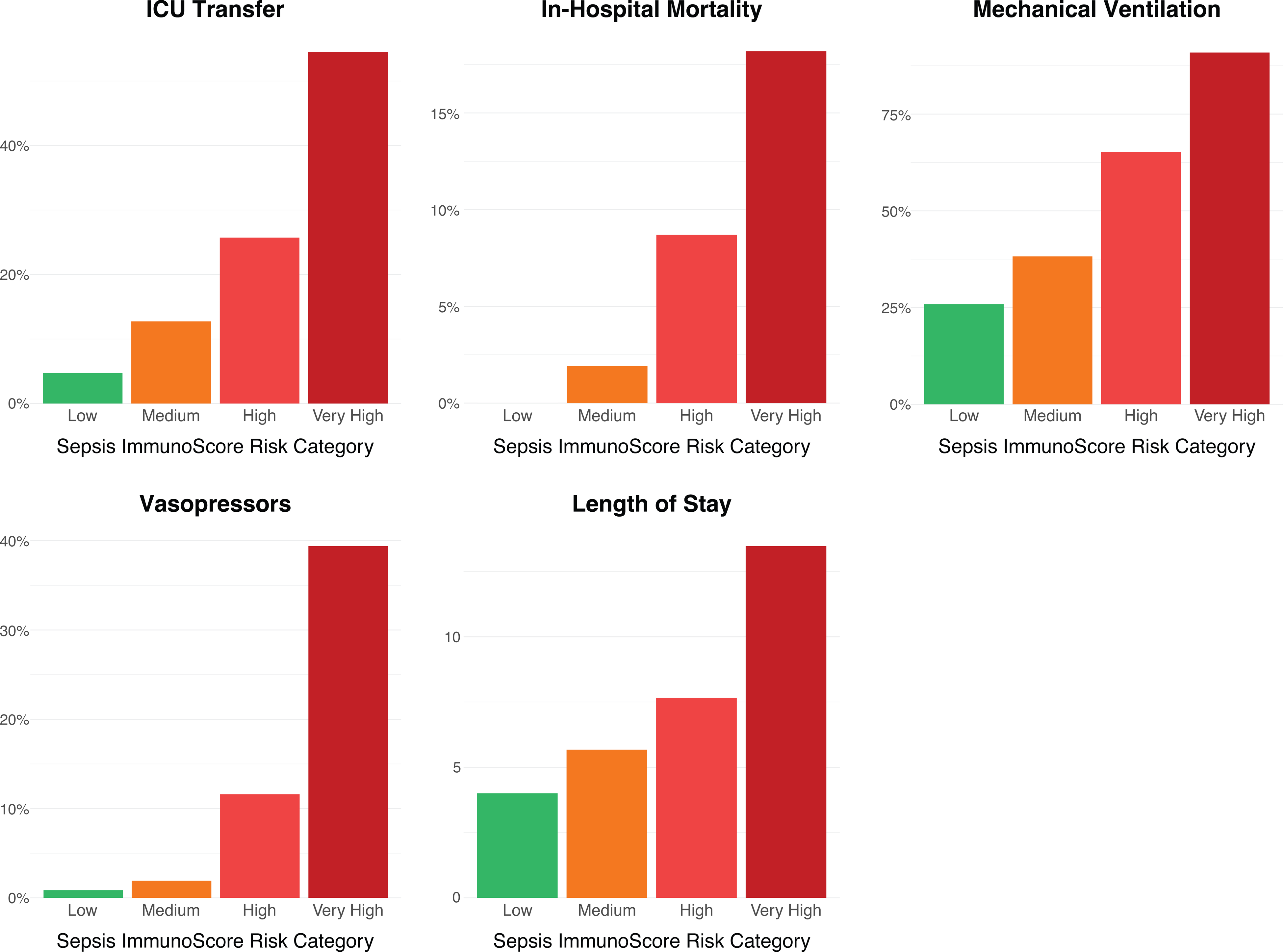
Sepsis ImmunoScore Risk Stratification for Morbidity and Mortality (External Validation). Barplots are shown for the derivation, internal validation, and external validation datasets for the secondary endpoints P.V.s (ICU transfer within 24 hours, in-hospital mortality, mechanical ventilation within 24 Hours, vasopressor administration within 24 Hours, and length of stay from inclusion time) for each Sepsis ImmunoScore risk stratification category. Dashed lines indicate the 95% C.I.s.

**Table 4.** External Validation ImmunoScore Risk Stratification for Morbidity and Mortality.

| Secondary Outcome | Sepsis Risk Category | Total Patients | Patients with Event | Predictive Value [95% CI] | Likelihood Ratio [95% CI] | Days [95% CI] | Cochran-Armitage Test (p-value) |
| --- | --- | --- | --- | --- | --- | --- | --- |
| ICU Transfer within 24 Hours | Low | 232 | 11 | 4.7% [2.4%, 8.3%] | 0.2 [0.1, 0.4] | - | < 0.001 |
|  | Medium | 157 | 20 | 12.7% [8.0%, 19.0%] | 0.7 [0.4, 1.1] | - |  |
|  | High | 276 | 71 | 25.7% [20.7%, 31.3%] | 1.7 [1.3, 2.1] | - |  |
|  | Very High | 33 | 18 | 54.6% [36.4%, 71.9%] | 5.8 [3.0, 11.3] | - |  |
| In-Hospital Mortality | Low | 232 | 0 | 0.0% [0.0%, 1.6%] | 0.0 [0.0, -] | - | < 0.001 |
|  | Medium | 157 | 3 | 1.9% [0.4%, 5.5%] | 0.4 [0.1, 1.2] | - |  |
|  | High | 276 | 24 | 8.7% [5.7%, 12.7%] | 1.9 [1.3, 2.9] | - |  |
|  | Very High | 33 | 6 | 18.2% [7.0%, 35.5%] | 4.5 [1.9, 10.7] | - |  |
| Mechanical Ventilation within 24 Hours | Low | 232 | 6 | 2.6% [1.0%, 5.5%] | 0.5 [0.2, 1.2] | - | 0.008 |
|  | Medium | 157 | 6 | 3.8% [1.4%, 8.1%] | 0.8 [0.4, 1.8] | - |  |
|  | High | 276 | 18 | 6.5% [3.9%, 10.1%] | 1.4 [0.9, 2.2] | - |  |
|  | Very High | 33 | 3 | 9.1% [1.9%, 24.3%] | 2.0 [0.6, 6.6] | - |  |
| Vasopressor<br>within 24 Hours | Low | 232 | 2 | 0.9% [0.1%, 3.1%] | 0.1 [0.0, 0.5] | - | < 0.001 |
|  | Medium | 157 | 3 | 1.9% [0.4%, 5.5%] | 0.3 [0.1, 0.8] | - |  |
|  | High | 276 | 32 | 11.6% [8.1%, 16.0%] | 1.7 [1.2, 2.4] | - |  |
|  | Very High | 33 | 13 | 39.4% [22.9%, 57.9%] | 8.4 [4.2, 16.7] | - |  |
| Length of Stay | Low | 232 | 232 | - | - | 4.0 [3.5, 4.9] |  |
|  | Medium | 157 | 157 | - | - | 5.7 [4.9, 7.0] |  |
|  | High | 276 | 276 | - | - | 7.7 [6.5, 8.5] |  |
|  | Very High | 33 | 33 | - | - | 13.5 [7.1, 19.1] |  |

### Sepsis-3 Diagnostic and Prognostic Sensitivity Analysis

As a secondary sensitivity analysis, we also analyzed the performance of the Sepsis ImmunoScore using alternative definitions of the sepsis outcome based on the timing of sepsis. Namely, to assess **diagnostic performance**, we analyzed the Sepsis ImmunoScore’s ability to discriminate patients who had sepsis at initial evaluation (n=99; 15.3%) from those who never had sepsis within 24 hours (n=547, 84.7%). We also assessed **prognostic performance** by analyzing the Sepsis ImmunoScore’s ability to discriminate patients without sepsis at initial evaluation who later develop sepsis within 24 hours (n=52; 7.4%) from those who never had sepsis (n=547, 92.6%). The AUC for sepsis diagnostic performance in the external validation set was 0.84 (0.78-0.89), and for the prognostic performance, it was 0.76 (0.68 – 0.84). The Sepsis ImmunoScore risk categories demonstrated similar results to the original combined analysis for both the diagnosis (**Table S4**) and prognosis of sepsis (**Table S5**). These data support good diagnostic and prognostic performance of the Sepsis ImmunoScore in this secondary analysis.

## Discussion

The Sepsis ImmunoScore is a comprehensive, multidimensional AI/ML tool that combines demographics, vital signs, clinical laboratory tests, and sepsis focused laboratory tests to assess risk of sepsis and risk of adverse outcomes. In this study, we developed the Sepsis ImmunoScore and evaluated its ability to serve as a risk-stratification tool for patients with suspected infection, and its ability to predict the diagnosis of sepsis and adverse clinical outcomes. We found the Sepsis ImmunoScore highly predictive of sepsis and secondary outcomes of in-hospital mortality, hospital length of stay, ICU admission, mechanical ventilation, and vasopressor administration within 24 hours. We also found that the Sepsis Immunoscore was predictive in diagnostic (predicting if a patient has sepsis at initial evaluation) and prognostic (predicting if a patient without sepsis at initial evaluation will develop sepsis) approaches.

There are a number of FDA-approved diagnostic tools available for patients with an infection; however, they are typically in the form of a single blood biomarker or sometimes multiple blood biomarkers. Procalcitonin is a biomarker that evaluates the risk of progression to severe sepsis and septic shock in critically ill patients upon their first day in the ICU.^21–25^ The IntelliSep Test is a blood test that measures leukocyte biophysical properties to create a score that identifies sepsis with organ dysfunction manifesting within the first three days after testing for adult patients with signs and symptoms of infection who present to the emergency department.^26–28^ Another test by Beckman, the Coulter Cellular Analysis System’s Early Sepsis Indicator measures Monocyte Distribution Width to identify sepsis risk.^29–31^ Other tests distinguish bacterial from non-bacterial infection in the E.D. or urgent care settings such as the FebriDx test which measures myxovirus resistance protein A and CRP from finger-stick blood.^32–35^ The MeMed BVTM measures blood concentrations of TRAIL, IP-10, and CRP to also distinguish patients with bacterial infections from those without.^36–39^ The Sepsis ImmunoScore uses multidimensional inputs across different domains (demographics, vital signs, laboratory tests etc.) plus sepsis biomarkers to create a comprehensive risk score for a given individual. The intent of the ImmunoScore is to embed in an EMR so that it can pull the different requisite inputs and display the score when it is ordered as a diagnostic test.

The Sepsis ImmunoScore was created with the intention of serving as an adjunct to clinical decision-making utilizing a Bayesian approach so that the clinician may combine the results of the Sepsis ImmunoScore with clinical assessment and traditional testing to make clinical decisions. Rather than relying on a single cutoff to classify results as “normal” or “abnormal,” we report four risk bands to capture a more similar test performance over a narrower range of values. For instance, approximately 1/3 of patients test in the low-risk band, which has a likelihood ratio of 0.1, a sepsis prevalence of 3%, and a sensitivity of 95%. This low-risk band can help “rule-out” sepsis in a patient with low to moderate clinical risk, assisting decisions for outpatient management. Conversely, for patients with moderate to high clinical risk, a high or very high Sepsis ImmunoScore is helpful from a Bayesian approach as a “rule-in” test. We also note that the Sepsis ImmunoScore bands are associated with the use of critical interventions such as mechanical ventilation, vasopressor use, and mortality; thus, this may also inform clinicians when making clinical decisions such as patient disposition. When the clinical assessment is disparate from the Sepsis ImmunoScore, additional observations, assessments, and possibly testing may be warranted.

While no other AI/ML tools are FDA authorized for sepsis, many have been developed and clinically deployed, especially early detection tools that passively monitor patient data and alert clinicians when sepsis is suspected. The reported performance of these tools varies widely, and recent validation studies have raised concerns about their use.^40–43^ A large, external validation study of the widely deployed Epic Sepsis Model in 2021 reported an AUC of only 0.63,^40^ and recent reviews of validation studies of the Targeted Real-time Early Warning System (TREWS) score have raised concerns regarding the control group and false positives.^41^ Concerns of alert fatigue have also been raised for these systems, which may undermine their clinical utility.^44–46^ The Sepsis ImmunoScore differs from early-warning systems in that it is intended to be coupled with a clinical suspicion of infection (e.g. ordering of a blood culture) as opposed to implementation as a screening tool without specific context. However, it is still prone to misclassification in the clinical setting.

The application of AI/ML to medicine has great potential, much of which is underdeveloped in medicine. The Sepsis ImmunoScore used clinically available data reflective of patient biologic state and machine learning to incorporate and identify objective patient assessments that are causally related to sepsis and associated adverse outcomes. Input features were carefully curated to select for measures of patient biology and pathophysiology that underlie critical illness and are routinely collected or available in the setting of infection.^47^ We did not include as eligible covariates subjective determinations or interventions that could be heavily influenced by site-specific protocols, clinician-specific perspectives, or other peculiarities of care. In addition to accurately diagnosing sepsis in an external validation set, we attribute the simultaneous association of the Sepsis ImmunoScore with other adverse outcomes in part to an explicit focus on patient host response biology. The result of this careful synthesis is a diagnostic tool that capitalizes on the synergy of thoughtfully applied AI/ML to expertly curated biologic data to better equip—not replace—clinicians in their challenging fight against sepsis. Furthermore, while the current FDA authorization does not allow local calibration or model adjustment, it is to consider future efforts that, with changes in FDA regulation, could leverage the ability of AI/ML algorithms to locally calibrate to improve performance.

Sepsis represents an ongoing diagnostic challenge to clinicians due to its often subtle and heterogeneous presentation. Assessing the presence or risk of progression to sepsis, and the severity with associated clinical needs represents a continuing challenge to clinicians. The Sepsis ImmunoScore is unique in its approach due to its machine-learning based incorporation of 22 parameters to comprehensively assess a patient’s risk of being diagnosed with sepsis, plus its association with adverse outcomes. The ImmunoScore could serve as an adjunctive test to assist clinical decision making in the acute setting. Given its strong predictive ability, the Sepsis ImmunoScore has the potential to improve patient outcomes by informing physician decisions for patients potentially requiring sepsis-related care, such as the rapid administration of broad-spectrum antimicrobials, escalation of care, and administration of fluid or vasopressor medications. It also has the potential to help to reduce over-triage by more accurately identifying patients at low risk for deterioration due to infection, for example potentially allowing emergency department physicians to treat these low-risk patients in the outpatient setting and promote antimicrobial stewardship.

### Limitations

There are a number of limitations to our study. First, because we used 5 hospitals in the study, it is possible that our findings may not generalize to specific populations that may differ from our hospitals. Second, we relied upon an EMR extraction, so it is possible that missingness or the use of ICD10 codes may have led to misclassification of certain elements such as comorbidities. Third, this was an observational study so we cannot assess the impact of the ImmunoScore on clinical decision-making and changes in therapeutic approaches. Fourth, the primary outcome of Sepsis-3 within 24 hours relied upon an automated calculation in the derivation set and adjudication for presence of infection in the internal and external validation; thus, misclassification of outcome may have occurred. Fifth, our inclusion criteria used the ordering of a blood culture as a surrogate indicator for a clinical suspicion of infection and patients where there was a clinical suspicion may not have had a blood culture ordered or other patients may have had a blood culture performed who had a very low (or no) suspicion of infection. Sixth, we note that approximately 7.2% of the potentially eligible patients were excluded due to not having labs or vital signs available to calculate the Sepsis ImmunoScore. Since patients without the requisite inputs had a higher prevalence of being from the in-hospital or ICU setting (Table S6), it is possible this could mildly (given the low prevalence) effect the generalizability of the score. Finally, covariate missingness may have affected algorithm performance.

## Conclusions

The Sepsis ImmunoScore has demonstrated robust risk assessment performance in derivation, internal, and external validation. Future work is warranted to further establish its generalizability to other settings. Finally, additional studies are warranted to assess the impact of the Sepsis ImmunoScore on clinical decision-making, sepsis care, and associated resource utilization and costs. These investigations are ongoing.

## Data Availability

All data produced in the present study are unavailable

## Authorship Contributions

Dr. Shapiro and Bhargava had full access to all of the data in the study and take responsibility for the integrity of the data and the accuracy of the data analysis.

*Study concept and design:* Bhargava, Lopez-Espina, Schmalz, Watson, Zhao, Zhu, Bashir, and Reddy Jr., and Shapiro

*Acquisition, analysis, and interpretation of data*: Bhargava, Lopez-Espina, Schmalz, Khan, Urdiales, Updike, Kurtzman, Dagan, Doodlesack, Stenson, Sarma, Reseland, Lee, Kravitz, Antkowiak, Shvilkina, Espinosa, Halalau, Demarco, Davila, Davila, Sims, Maddens, Berghea, Smith, Palagiri, Ezekiel, Sadaka, Iyer, Crisp, Azad, Oke, Friederich, Syed, Gosai, Chawla, Evans, Thomas, Malkani, Patel, Mayer, Ali, Raghavakurup, Tafa, Singh, and Raouf

*Drafting of the manuscript:* Bhargava, Watson, and Shapiro

*Critical revision of the manuscript for important intellectual content*: All authors

*Statistical analysis:* Bhargava, Schmalz, and Watson

*Obtained funding:* Lopez-Espina, Watson, Bashir, and Reddy Jr.

*Administrative, technical, or material support:* Reddy Jr.

*Study supervision:* Bhargava, Lopez-Espina, Schmalz, Reddy Jr., and Shapiro

## Acknowledgement Statement

We are indebted to the study coordinators, research staff, and lab technicians who participated in the study. These contributions were part of these individuals’ jobs, and they did not receive additional compensation.

