## Supplementary material for "Development and Validation of the First FDA Authorized Artificial Intelligence/Machine Learning Diagnostic Tool for the Prediction of Sepsis Risk": Online Supplement

### Development and Validation of a Machine Learning Diagnostic Tool for the Prediction of Sepsis and Critical Illness Online Supplement

#### Study Population

The derivation cohort consisted of 2,366 participants enrolled between April 2017 and January 2021 from 3 hospitals: Carle Foundation Hospital (Urbana, IL), Mercy Hospital St. Louis (St. Louis, MO), and OSF Saint Francis Medical Center (Peoria, IL). The internal validation cohort consisted of 393 participants enrolled between February 2021 and June 2021 from 2 different hospitals: Mercy Hospital St. Louis (St. Louis, MO) and Our Saint Francis (Peoria, IL). The final external validation cohort consisted of 698 participants enrolled between January 2021 and July 2022 from 3 hospitals: William Beaumont University Hospital (Royal Oak, MI), Jesse Brown Department of Veteran Affairs Medical Center (Chicago, IL), and Beth Israel Deaconess Medical Center (Boston, MA).

#### Study Outcomes

##### Endpoints

Infection was assessed as present if any of the following criteria were met: 4 or more qualifying antimicrobial days (as recommended by the Centers for Disease Control and Prevention's QAD guidance), or evidence of a definite infection via culture results or other laboratory microbiological testing (excluding common contaminants). Organ dysfunction was considered present if there was a change in SOFA score of 2 or greater from baseline. If available, we calculated SOFA at baseline using historical laboratory measurements and diagnoses; otherwise, we assumed a value of 0. Clinical adjudication was performed for each subject in the internal and external validation cohorts using an independent review by three physicians blinded to each other's review and the Sepsis ImmunoScore result. Each adjudicator used the entirety of the patient's EMR to assess for the presence of infection and organ dysfunction to determine if the Sepsis-3 criteria were met. The assessment of organ dysfunction was based on whether the adjudicator observed an increase from baseline of the SOFA score of 2 points or more or identified other clinical evidence indicating the presence of life-threatening organ dysfunction. A clinically adjudicated final diagnosis of sepsis-3 within 24 hours was assigned to subjects for whom at least 2 adjudicators indicated that the subject had organ dysfunction caused by an infection within 24 hours of study inclusion.

#### Data Collection

##### C-Reactive Protein and Procalcitonin Measurements

We posited a priori that C-reactive protein (CRP) and procalcitonin (PCT) would be valuable components of a risk prediction score, so we collected discarded specimens for external testing. We defined eligible samples as plasma collected from the patient in a Li-Hep tube within a 6-hour window centered at the patient's first blood culture order. Upon identification, the site's lab technicians processed the sample by spinning, aliquoting them into multiple 175-microliter tubes, and placing them into -80$^{o}$ C freezer. The institutions then shipped the frozen aliquots to our central laboratory, where we stored them in a -80$^{o}$ C freezer. CRP and PCT measurements in the derivation cohort were obtained by thawing and measuring the frozen samples using the Luminex Assay Platform. CRP and PCT measurements in the internal and validation cohort were conducted by one off three CLIA-certified laboratories using a Cobas 8000 Analyzer Series Module e801 and Cobas 6000 Analyzer Series Module c501 for PCT and CRP concentrations, respectively. All measurements were obtained within 1 year of sample freezing.

#### Sepsis ImmunoScore

##### Risk Category Development

The boundary between the low and medium risk categories was set at 12.2, the maximally sensitive risk score with a false positive rate no greater than 50%; the boundary between the medium and high risk categories was set at 30.6, the point on the receiver operating characteristic (ROC) curve closest in Euclidian distance to (0,1); and the boundary between the high and very high risk categories was set at 87.2, the risk score below which 95% of the out-of-bag predictions fell.

##### Tables

##### Table S1. AUROC of Procalcitonin and Sepsis ImmunoScore for Sepsis in All Cohorts

| Cohort | Procalcitonin AUROC (95% CI) | C-reactive Protein AUROC (95% CI) | Sepsis ImmunoScore AUROC (95% CI) |
|---|---|---|---|
| Derivation | 0.71 (0.69, 0.74) | 0.60 (0.58, 0.63) | 0.85 (0.83, 0.87) |
| Internal Validation | 0.71 (0.65, 0.77) | 0.61 (0.55, 0.67) | 0.80 (0.74, 0.86) |
| External Validation | 0.75 (0.70, 0.80) | 0.68 (0.63, 0.73) | 0.81 (0.77, 0.86) |

##### Table S2. Derivation and Internal Validation Sepsis ImmunoScore Risk Stratification for Sepsis

| Cohort | ImmunoScore Risk Category | Total Patients (N) | Septic Patients (N) | Sepsis PV [95% CI] | Sepsis Likelihood Ratio [95% CI] | Cochran-Armitage (*p*-value) |
|---|---|---|---|---|---|---|
| Derivation | Low | 849 | 47 | 5.5% [4.1%, 7.3%] | 0.1 [0.1, 0.2] | < 0.001 |
| (N = 2366) | Medium | 556 | 122 | 21.9% [18.6%, 25.6%] | 0.6 [0.5, 0.7] | |
| | High | 843 | 485 | 57.5% [54.1%, 60.9%] | 2.9 [2.6, 3.2] | |
| | Very High | 118 | 109 | 92.4% [86.0%, 96.5%] | 25.4 [13.0, 49.9] | |
| Internal | Low | 144 | 13 | 9.0% [4.9%, 14.9%] | 0.3 [0.2, 0.4] | < 0.001 |
| Validation | Medium | 91 | 15 | 16.5% [9.5%, 25.7%] | 0.5 [0.3, 0.9] | |
| (N = 393) | High | 141 | 66 | 46.8% [38.4%, 55.4%] | 2.3 [1.8, 3.0] | |
| | Very High | 17 | 14 | 82.4% [56.6%, 96.2%] | 12.3 [3.6, 42] | |

##### Table S3. Derivation and Internal Validation Sepsis ImmunoScore Risk Stratification for Morbidity and Mortality

| Cohort | Secondary Outcome | Sepsis Risk Category | Total Patients | Patients with Event | Predictive Value [95% CI] | Likelihood Ratio [95% CI] | Days [95% CI] | Cochran-Armitage |
|---|---|---|---|---|---|---|---|---|

| | | | | | | | | p-value |
|---|---|---|---|---|---|---|---|---|
| **Derivation** | ICU Transfer within 24 Hrs[1] | Low | 624 | 43 | 6.9% [5.03%, 9.2%] | 0.2 [0.2, 0.3] | - | < 0.001 |
| | | Medium | 426 | 76 | 17.8% [14.3%, 21.8%] | 0.7 [0.54, 0.9] | - | |
| | | High | 625 | 249 | 39.2% [35.4%, 43.1%] | 2.0 [1.76, 2.3] | - | |
| | | Very High | 88 | 60 | 68.2% [57.4%, 77.7%] | 6.7 [4.32, 10.5] | - | |
| | In-Hospital Mortality | Low | 849 | 6 | 0.7% [0.3%, 1.5%] | 0.1 [0.05, 0.2] | - | < 0.001 |
| | | Medium | 556 | 20 | 3.6% [2.2%, 5.5%] | 0.6 [0.4, 0.9] | - | |
| | | High | 843 | 87 | 10.3% [8.4%, 12.6%] | 1.7 [1.4, 2.] | - | |
| | | Very High | 118 | 34 | 28.8% [20.9%, 37.9%] | 6.1 [4.1, 9.0] | - | |
| | Mechanical Ventilation within 24 Hrs | Low | 849 | 9 | 1.1% [0.5%, 2.00%] | 0.2 [0.1, 0.4] | - | < 0.001 |
| | | Medium | 556 | 16 | 2.9% [1.7%, 4.63%] | 0.5 [0.3, 0.8] | - | |
| | | High | 843 | 70 | 8.3% [6.5%, 10.37%] | 1.5 [1.2, 2.0] | - | |
| | | Very High | 118 | 36 | 30.5% [22.4%, 39.7%] | 7.5 [5.1, 11.0] | - | |
| | Vasopressor within 24 Hrs | Low | 849 | 3 | 0.4% [0.1%, 1.0%] | 0.1 [0.0, 0.2] | - | < 0.001 |
| | | Medium | 556 | 12 | 2.2% [1.1%, 3.7%] | 0.3 [0.2, 0.5] | - | |
| | | High | 843 | 92 | 10.9% [8.9%, 13.2%] | 1.8 [1.4, 2.2] | - | |
| | | Very High | 118 | 45 | 38.1% [29.4%, 47.5%] | 9.0 6.2, 12.9] | - | |
| | Length of Stay[2] | Low | 849 | 849 | - | - | 3.1 [2.9, 3.3] | |
| | | Medium | 556 | 556 | - | - | 4.3 [4.0, 4.8] | |
| | | High | 843 | 843 | - | - | 6.7 [6.2, 7.0] | |
| | | Very High | 118 | 118 | - | - | 14.6 [10.7, 21.0] | |
| **Internal Validation** | ICU Transfer within 24 Hrs | Low | 144 | 14 | 9.7% [5.4%, 15.8%] | 0.22 [0.1, 0.4] | - | < 0.001 |
| | | Medium | 91 | 29 | 31.9% [22.5%, 42.5%] | 1.0 [0.6, 1.4] | - | |
| | | High | 141 | 73 | 51.8% [43.2%, 60.3%] | 2.2 [1.6, 2.9] | - | |
| | | Very High | 17 | 14 | 82.4% [56.6%, 96.2%] | 9.4 [2.7, 32.6] | - | |
| | In-Hospital Mortality | Low | 144 | 1 | 0.% [0.00%, 3.81%] | 0.1 [0.0, 0.5] | - | < 0.001 |
| | | Medium | 91 | 6 | 6.6% [2.5%, 13.8%] | 0.8 [0.3, 1.7] | - | |
| | | High | 141 | 19 | 13.5% [8.3%, 20.2%] | 1.7 [1.1, 2.7] | - | |
| | | Very High | 17 | 7 | 41.2% [18.4%, 67.1%] | 7.6 [3.0, 20.0] | - | |
| | Mechanical Ventilation within 24 Hrs | Low | 144 | 2 | 1.4% [0.2%, 4.9%] | 0.2 [0.0, 0.7] | - | < 0.001 |
| | | Medium | 91 | 5 | 5.5% [1.8%, 12.4%] | 0.7 [0.3, 1.8] | - | |
| | | High | 141 | 16 | 11.4% [6.6%, 17.8%] | 1.6 [1.0, 2.7] | - | |
| | | Very High | 17 | 6 | 35.3% [14.2%, 61.7%] | 6.9 [2.6, 18.3] | - | |
| | Vasopressor within 24 Hrs | Low | 144 | 2 | 1.4% [0.2%, 4.9%] | 0.1 [0.0, 0.6] | - | < 0.001 |
| | | Medium | 91 | 4 | 4.4% [1.2%, 10.9%] | 0.5 [0.2, 1.2] | - | |
| | | High | 141 | 20 | 14.2% [8.9%, 21.1%] | 1.6 [1.04, 2.6] | - | |
| | | Very High | 17 | 10 | 58.8% [32.9%, 81.6%] | 14.2 [5.5, 36.8] | - | |
| | Length of Stay[2] | Low | 144 | 144 | - | - | 3.3 [3.0, 4.0] | |
| | | Medium | 91 | 91 | - | - | 5.9 [4.6, 7.4] | |
| | | High | 141 | 141 | - | - | 7.1 [5.8, 9.8] | |
| | | Very High | 17 | 17 | - | - | 19.4 [7.9, -] | |

[1] Hospital Location from Carle Foundation Hospital was unavailable

[2] Subjects with an in-hospital mortality were treated with an infinite length of stay due to competing risk

#### Table S4. Sepsis ImmunoScore Performance for Subjects Meeting Sepsis-3 at Evaluation Time (Diagnostic Sepsis and No Sepsis)

| Risk Group | Septic Patients (N) | Total Patients (N) | PV (95% CI) | SSLR (95% CI) |
|---|---|---|---|---|

| | | | | |
|---|---|---|---|---|
| Low | 2 | 227 | 0.9% [0.1%, 3.2%] | 0.1 [0.0, 0.2] |
| Medium | 12 | 149 | 8.0% [4.2%, 13.7%] | 0.5 [0.3, 0.9] |
| High | 72 | 247 | 29.2% [23.6%, 35.3%] | 2.3 [1.8, 2.9] |
| Very High | 13 | 23 | 56.5% [34.5%, 76.8%] | 7.2 [3.2, 16.2] |

**Table S5. Sepsis ImmunoScore Performance for Subjects Meeting Sepsis-3 After Evaluation Time (Prognostic Sepsis and No Sepsis)**

| Risk Group | Septic Patients (N) | Total Patients (N) | PV (95% CI) | SSLR (95% CI) |
|---|---|---|---|---|
| Low | 5 | 230 | 2.2% [0.7%, 5.0%] | 0.2 [0.1, 0.6] |
| Medium | 8 | 145 | 5.5% [2.4%, 10.6%] | 0.6 [0.3, 1.2] |
| High | 29 | 204 | 14.2% [9.7%, 19.8%] | 1.7 [1.2, 2.5] |
| Very High | 10 | 20 | 50.0% [27.2%, 72.8%] | 10.5 [4.4, 25.0] |

**Table S6. Demographic Comparison of Subjects with an Invalid Sepsis ImmunoScore Results vs Valid Sepsis ImmunoScore Result**

| Characteristic | Patients Encounters, No. (%) | |
|---|---|---|
| | Invalid Score (N = 269) | Valid Score (N = 3457) |
| Clinical Site | | |
| Beth Israel Deaconess Medical Center – Boston, MA | 14 (5.2) | 356 (10.3) |
| OSF – Peoria, IL | 85 (31.6) | 799 (23.1) |
| Jesse Brown VA - Chicago, IL | 8 (3.0) | 65 (1.9) |
| Mercy Health - St. Louis, MO | 74 (27.5) | 1367 (39.5) |
| Beaumont - Royal Oak, MI | 26 (9.7) | 277 (8.0) |
| Carle Foundation Hospital - Urbana, IL | 62 (23.0) | 593 (17.2) |
| Age (mean (SD)) | 65.00 [53.00, 75.00] | 66.00 [53.00, 76.00] |
| Male (%) | 142 (52.8) | 1796 (52.0) |
| Race | | |
| American Indian or Alaska Native | 0 (0.0) | 3 (0.1) |
| Asian | 3 (1.1) | 28 (0.8) |
| Black or African American | 50 (18.6) | 526 (15.2) |
| Native Hawaiian or Other Pacific Islander | 0 (0.0) | 1 (0.0) |
| Unknown | 17 (6.3) | 216 (6.2) |
| White | 199 (74.0) | 2683 (77.6) |
| Ethnicity | | |
| Hispanic or Latino | 6 (2.2) | 124 (3.6) |

| | | |
|---|---|---|
| Not Hispanic or Latino | 196 (72.9) | 2674 (77.4) |
| Unknown | 67 (24.9) | 659 (19.1) |
| High-Risk Comorbidities | | |
| Acute Myocardial Infarction (%) | 18 (6.7) | 151 (4.4) |
| History of Myocardial Infarction (%) | 15 (5.6) | 176 (5.1) |
| Congestive Heart Failure (%) | 74 (27.5) | 856 (24.8) |
| Peripheral Vascular Disease (%) | 27 (10.0) | 346 (10.0) |
| Cerebrovascular Disease (%) | 17 (6.3) | 233 (6.7) |
| Chronic Obstructive Pulmonary Disease (%) | 63 (23.4) | 884 (25.6) |
| Dementia (%) | 13 (4.8) | 284 (8.2) |
| Paralysis (%) | 12 (4.5) | 98 (2.8) |
| Diabetes (%) | 70 (26.0) | 887 (25.7) |
| Diabetes with Complications (%) | 52 (19.3) | 671 (19.4) |
| Renal Disease (%) | 78 (29.0) | 998 (28.9) |
| Mild Liver Disease (%) | 20 (7.4) | 231 (6.7) |
| Moderate and Severe Liver Disease (%) | 5 (1.9) | 106 (3.1) |
| Peptic Ulcer Disease (%) | 7 (2.6) | 66 (1.9) |
| Rheumatologic Disease (%) | 9 (3.3) | 155 (4.5) |
| AIDS (%) | 3 (1.1) | 25 (0.7) |
| Immunocompromised (%) | 72 (26.8) | 777 (22.5) |
| COVID-19 (%) | 18 (6.7) | 290 (8.4) |
| Blood Culture Setting (%) | | |
| Critical Care | 25 (9.3) | 189 (5.5) |
| Emergency Department | 114 (42.4) | 2291 (66.3) |
| Non-Critical Inpatient | 66 (24.5) | 362 (10.5) |
| Non-ED Short Stay | 2 (0.7) | 22 (0.6) |
| Unknown[1] | 62 (23.0) | 593 (17.2) |

[1] Hospital Location data was not provided for Carle Foundation Hospital Subjects

#### Figures

##### Figure S1. Flow Chart. For Selection of Subjects and Exclusion of Ineligible Subjects

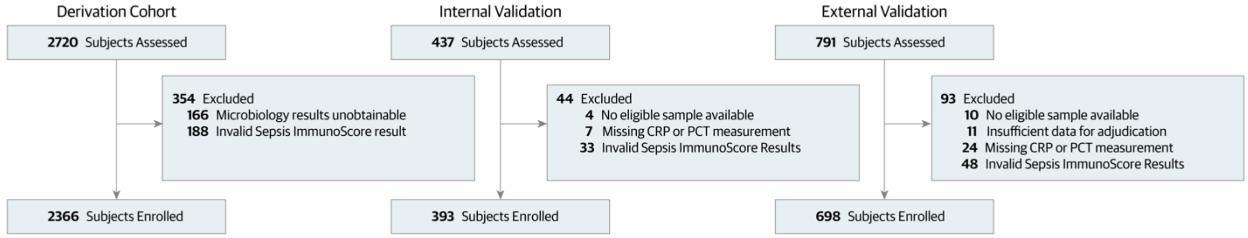

This waterfall diagram illustrates the enrollment process for adult subjects (≥18 years old) suspected of infection (based on a blood culture order) who presented to the emergency department (ED) or hospital across the derivation, internal validation, and external validation cohorts. Several exclusion criteria were applied between subject assessment and enrollment. These included: Microbiology results unobtainable for a subset of patients in the derivation dataset which may have altered the automated Sepsis-3 label, Invalid Sepsis ImmunoScore results due to missing laboratory data (measurements for WBC, platelet count, creatinine, or BUN within a timeframe spanning 24 hours before the blood culture order [study entry] to 3.5 hours after), Invalid Sepsis ImmunoScore results due to missing vital signs (data for systolic BP, diastolic BP, SpO2, heart rate, or respiratory rate between six hours before study entry and 3.5 hours after), inability to obtain a PCT or CRP measurement due to potential sample carry over or sample volume constraints, inability to collect a lithium-heparin blood sample within a 6-hour window around study entry, and insufficient data for proper adjudication.

**Figure S2. Sepsis ImmunoScore Risk Stratification for Morbidity and Mortality: Time to Event (External Validation Cohort)**

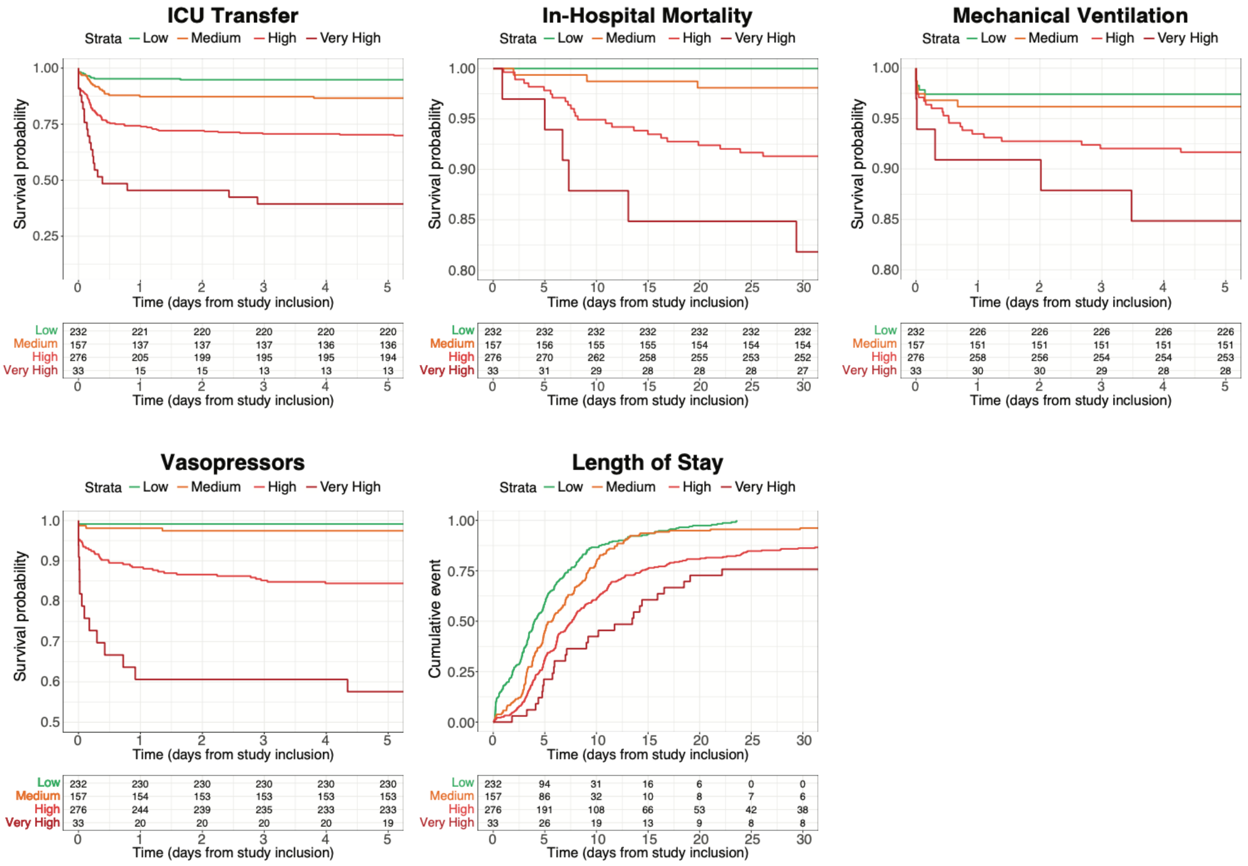

Cumulative Incidence Plots for are shown for external validation datasets for the secondary endpoints (ICU transfer, in-hospital mortality, mechanical ventilation, vasopressor administration, and length of stay from inclusion time) for each Sepsis ImmunoScore risk stratification category. All subjects who died in-hospital were assumed to have a length of stay > 30 days. All subjects who were discharged and did not die in-hospital were assumed to have an in-hospital mortality time > 30 days. All subjects who were discharged or died prior their first ICU transfer, mechanical ventilation, or vasopressor administration were assumed to have an event time of > 5 days.
